# Protection of hybrid immunity against SARS-CoV-2 reinfection and severe COVID-19 during periods of Omicron variant predominance in Mexico

**DOI:** 10.1101/2022.12.02.22282981

**Authors:** José Antonio Montes-González, Christian Arturo Zaragoza-Jiménez, Neftali Eduardo Antonio-Villa, Carlos A. Fermín-Martínez, Daniel Ramírez-García, Arsenio Vargas-Vázquez, Rosaura Idania Gutiérrez-Vargas, Gabriel García-Rodríguez, Hugo López-Gatell, Sergio Iván Valdés-Ferrer, Omar Yaxmehen Bello-Chavolla

## Abstract

**BACKGROUND:** With widespread transmission of the Omicron SARS-CoV-2 variant, reinfections have become increasingly common. Here, we explored the role hybrid immunity, primary infection severity, and variant predominance on the risk of reinfection and severe COVID-19 during periods of Omicron predominance in Mexico.

**METHODS:** We analyzed reinfections in Mexico in individuals with ≥90 days from a previous primary infection using a national surveillance registry of SARS-CoV-2 cases from March 3^rd^, 2020, until August 13^th^, 2022. Immunity-generating events included primary infection, partial or full vaccination and vaccine boosting. Reinfections were matched by age and sex with controls with primary SARS-CoV-2 infection and negative RT-PCR or antigen test ≥90 days after infection to explore risk factors for reinfection and reinfection-associated severe COVID-19. We also explored the protective role of heterologous vs. homologous vaccine boosters against reinfection or severe COVID-19 in fully vaccinated individuals.

**RESULTS:** We detected 231,202 SARS-CoV-2 reinfections in Mexico, with most occurring in unvaccinated individuals (41.55%). Over 207,623 reinfections occurred during periods of Omicron (89.8%), BA.1 (36.74%) and BA.5 (33.67%) subvariant predominance and a case-fatality rate of 0.22%. Vaccination protected against reinfection, without significant influence of the order of immunity-generating events and provided >90% protection against severe reinfections. Heterologous booster schedules were associated with ∼11% and ∼54% lower risk for reinfection and reinfection-associated severe COVID-19 respectively, modified by time-elapsed since the last immunity-generating event.

**CONCLUSIONS:** SARS-CoV-2 reinfections have increased during periods of Omicron predominance. Hybrid immunity provides protection against reinfection and reinfection-associated severe COVID-19, with potential benefit from heterologous booster schemes.

**RESEARCH IN CONTEXT:** *Evidence before this study:* We searched PubMed for the terms “SARS-CoV-2” AND “reinfection” AND “hybrid immunity” until November 20^th^, 2022, and identified a few population studies previously conducted in Israel, Sweden, Qatar, United States and Canada which explored risk of reinfection and the protective role of hybrid immunity in individuals with one, two or three doses of COVID-19 vaccines, predominantly during periods of predominance of Omicron BA.1 and BA.2 subvariants. Notably, no studies were conducted in any Latin American country or reported on the benefit of heterologous booster schemes or the order of immunity-generating events.

*Added value of this study:* We report the results of nation-wide study in Mexico of over 230,000 SARS-CoV-2 reinfections, with ∼90% occurring during periods of Omicron predominance. We identified that vaccination provided additional benefit on reducing the risk of SARS-CoV-2 reinfection, with the highest benefit observed in individuals with complete vaccination and booster protocols prior to primary infection or with primary infection during periods of BA.1 and BA.2 subvariant predominance. Hybrid immunity also provides a substantial reduction in the risk of reinfection-associated severe COVID-19, with >90% reduction in risk compared to unvaccinated individuals with previous SARS-CoV-2 infection, regardless of the order of immunity-generating events. Finally, heterologous COVID-19 booster schedules were associated with ∼11% and ∼54% lower risk for reinfection and reinfection-associated severe COVID-19 respectively, modified by time-elapsed since the last immunity-generating event.

*Implications of all the available evidence:* Our results support that COVID-19 vaccination and boosters provide additional benefit to protect against SARS-CoV-2 reinfection and reinfection-associated severe COVID-19. The use of heterologous boosters appears to provide additional protection in previously infected individuals and such schemes may prove beneficial to increase vaccination coverage as newer, more transmissible variants emerge.

## INTRODUCTION

As the COVID-19 pandemic continues, an increasing number of previously infected individuals have become reinfected with SARS-CoV-2 (1). Evidence of SARS-CoV-2 reinfection was first documented in August 2020 (2), and was initially considered a rare event (1,3–7). With the emergence of the Omicron SARS-CoV-2 variant and its subvariants, shown to possess higher capacity for immune escape (8,9) and transmissibility (10), the largest increase of infection and reinfection rates has been documented(11). With Omicron, approximately 41% of the population in some countries are estimated to be at risk of SARS-CoV-2 reinfection (12). Although SARS-CoV-2 reinfections are described as less severe than primary infections (13), severe events continue to be reported despite increasing vaccination rates (3,14); furthermore, some studies have reported no difference in severity between prior infections and reinfections (15,16). For individuals who survive reinfections, a link with all-cause mortality and hospitalization in the acute and post-acute phase has been reported, as well as a relation between the frequency of COVID-19 reinfections and the prevalence of post-acute COVID-19 conditions(17).

Individuals with previous infection and at least one dose of a COVID-19 vaccine are benefitted with hybrid immunity, where natural immunity due to infection and vaccination-acquired immunity interact to enhance protection against infection and severe disease. Hybrid immunity in individuals vaccinated at some point before reinfection has been previously demonstrated (18–20). Previous data also supports an imprinting role on B-cell immune response from first exposure to COVID-19, either through vaccines or previous SARS-CoV-2 infection, which would condition the host response towards future infections (10,21,22). Despite increasing evidence of modifying factors of hybrid immunity such as time-dependent waning immunity, vaccine boosters, severity of primary infections (23) and circulating SARS-CoV-2 variants, real-world data on risk of reinfections with hybrid immunity, particularly for circulating Omicron subvariants is scarce (24). Here, we evaluated the role of primary infection and hybrid immunity on the risk of reinfection in individuals with a previous SARS-CoV-2 infection registered during periods of Omicron variant predominance in Mexico. We also evaluated the influence of the order of immunity-generating events, previous hospitalization, and SARS-CoV-2 variant predominance on the risk of reinfection and reinfection-associated severe outcomes using a nation-wide COVID-19 registry.

## METHODS

### Study population

We assessed cases of suspected SARS-CoV-2 reinfection using the SISVER registry, a daily updated nation-wide surveillance system of suspected SARS-CoV-2 cases in Mexico (25,26), managed by the General Directorate of Epidemiology of the Mexican Ministry of Health. Detailed sociodemographic and clinical information is ascertained, including details of SARS-CoV-2 infection and clinical course, as well as vaccination status, date and vaccine applied. For this analysis, from March 3^rd^, 2020, until August 13^th^, 2022, we analyzed survivors of a first confirmed SARS-CoV-2 infection with a second RT-PCR or antigen test for suspected SARS-CoV-2 reinfection ≥90 days after primary infection during periods of predominance of the Omicron variant. A flow chart of included and excluded subjects is presented in **Figure 1**.

**Figure 1.**
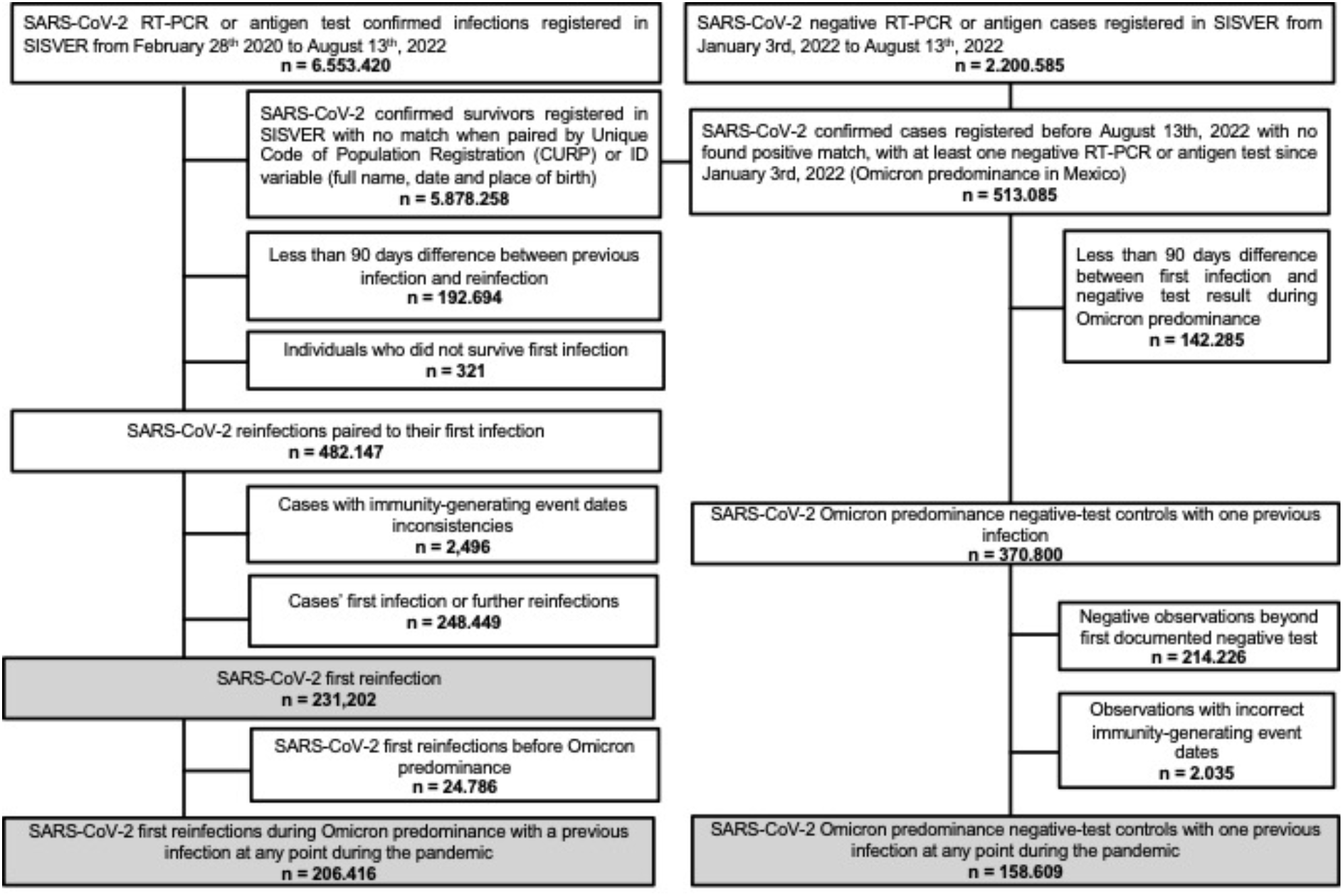
Flow chart of included and excluded subjects with SARS-CoV-2 documented reinfection or negative test as reported in SISVER from March 2020 to August 13^th^, 2022.

### Definition of immunity-generating events

An immunity-generating event was considered as either a vaccination against COVID-19 (first, second or booster shot) or a confirmed SARS-CoV-2 infection, either primary or reinfection. Reinfections were defined as a confirmed SARS-CoV-2 infection using RT-PCR or antigen test ≥90 days from confirmed primary SARS-CoV-2 infection (27). Nationally-available SARS-CoV-2 vaccines applied in Mexico during this period included BNT162b2, mRNA-12732, Gam-COVID-Vac, Ad5-nCoV, Ad26.COV2.S, ChAdOx1, NVX-CoV2373 and CoronaVac (28). Vaccination schedule for most vaccines considered two-doses, except for one-dose Ad5-nCoV and Ad26.COV2.S vaccines. Fully vaccinated individuals were considered if they had completed the vaccination schedule ≥14 days before the evaluated outcome (24,29). Partially vaccinated individuals were considered if they had one out of a two-dose vaccine schedule or if the outcome happened <14 days in an otherwise completely vaccinated individual. COVID-19 vaccines were categorized according to their platform. Booster vaccination was considered if fully vaccinated individuals received an additional shot of COVID-19 vaccine and ≥7 days had passed since vaccination(30), otherwise were reclassified as fully vaccinated. Booster schedules were categorized as homologous if booster shots were the same as the primary vaccination and heterologous if different.

### Determinants of reinfection and severe COVID-19 risk

Previous evidence has shown varying degrees of protection for SARS-CoV-2 reinfection and severe COVID-19 (defined by hospitalization, ICU admission, intubation, or death) based on the variant responsible for primary and second infection (31,32), time since vaccination or primary infection (32,33), and antibody response (34–36). Therefore, we evaluated the following variables:

1. *Predominant SARS-CoV-2 variant for first infection* – A fraction of COVID-19 samples are sequenced by authorized national laboratories and submitted to the GISAID platform. Infection was assumed to be most likely caused by the predominant variant based on date of symptom onset. Based on data submitted to GISAID(37), from March 3^rd^ 2020 until March 30^st^, 2021 the predominant SARS-CoV-2 variant was the ancestral strain, followed by predominance of the B.1.1.519 variant until June 6^th^, 2021, the P.1 (Gamma) variant until July 4^th^, 2021, the B.1.617.2 (Delta) variant until January 2nd, 2022. B.1.1.529.1 (Omicron) BA.1 subvariant was considered from January 3^rd^, 2022, until April 24^th^, 2002, followed by Omicron subvariant BA.2 until June 19^th^, 2022.
2. *Predominant SARS-CoV-2 variant for reinfection* – Categorized based on the date of symptom onset for the reinfections according to GISAID. Periods of Omicron subvariants BA.1 and BA.2 dominance were the same as previously described. Predominance of the BA.4 subvariant was considered from June 20^th^ until July 3^rd^, 2022, followed by BA.5 subvariant predominance until August 13^th^, 2022.
3. *Previous COVID-19-related hospitalization* – Defined if an individual was hospitalized and survived their primary SARS-CoV-2 infection.
4. *Order of immunity-generating events* – We developed an indicator variable which considered the order of immunity-generating events for a given individual, using as reference unvaccinated individuals with primary SARS-CoV-2 infection (**Supplementary Material**). This variable considered the order of first infection, partial, full or booster vaccination prior to their second SARS-CoV-2 testing for suspected reinfection.
5. *Time elapsed since last immunity-generating events* – Defined as time elapsed in months since the last immunity-generating event, either vaccination or infection. This was categorized based on whether individuals had ≥6 months since exposure, given previous evidence on vaccination or previous infection immunity waning over this period (38).

### Statistical analyses

#### Epidemiology of SARS-CoV-2 reinfections in Mexico

We characterized all SARS-CoV-2 reinfections in Mexico to explore sociodemographic and clinical characteristics. Reinfection incidence and mortality were calculated over the number of individuals who survived a confirmed primary SARS-CoV-2 infection. The number of confirmed reinfections was plotted over time and based on occurrence during periods of variant predominance in Mexico for the first and second SARS-CoV-2 infections using cross-tabulation matrix plots to visualize combinations of reinfections (**Supplementary Material**)

#### Determinants of reinfection and reinfection-associated severe COVID-19 risk

We matched SARS-CoV-2 reinfections to individuals with a negative SARS-CoV-2 test after primary infection using propensity score matching for age and sex. Next, we fitted conditional logistic regression models including matching weights to explore the role of previously defined determinants on the risk of reinfection. We also explored interaction effects between the order of immunity-generating events, and time elapsed since last exposure to an immunity-generating or hospitalization during primary SARS-CoV-2 infection. For models pertaining to risk of severe COVID-19 associated with reinfection, we only analyzed cases with confirmed reinfection and explored factors as described earlier.

#### Heterologous vs. homologous vaccine boosting

Given the diversity of COVID-19 vaccination schedules applied in Mexico, we explored risk of reinfection and severe COVID-19 associated with homologous and heterologous booster protocols, compared to fully vaccinated individuals, exploring similar determinants as described above using conditional logistic regression.

## RESULTS

### Epidemiology of SARS-CoV-2 reinfections in Mexico

We detected 231,202 confirmed reinfections over the study period, with most reinfections occurring during January and June in 2022. A steady rate of reinfection-associated mortality was observed starting from March 2021 (**Figure 2**). Most SARS-CoV-2 reinfections occurred in women (60.12%) and individuals aged 31-40 years (30.26% and a median time between reinfections of 362 days (IQR 196 – 531 days), **Supplementary Material**). Reinfections occurred primarily in unvaccinated individuals (41.55%) or cases in which primary infection occurred before completing vaccination schedules or receiving a booster shot (41.0%). Reinfections in individuals with comorbid diabetes or obesity were low (5.97% and 8.34%). Most primary infections occurred during predominance of ancestral strains (50.06%), followed by Delta (23.1%) and Omicron BA.1 (14.67%). Over 206,416 reinfections occurred during periods of predominance of the Omicron variant in Mexico (89.3%), mostly associated with predominance of Omicron BA.1 (36.74%) and Omicron BA.5 (33.67%) subvariants. We identified 3,261 hospitalizations related to reinfections (1.41%), and 515 deaths, with a reinfection fatality rate of 0.22% (**Table 1**).

**Figure 2.**
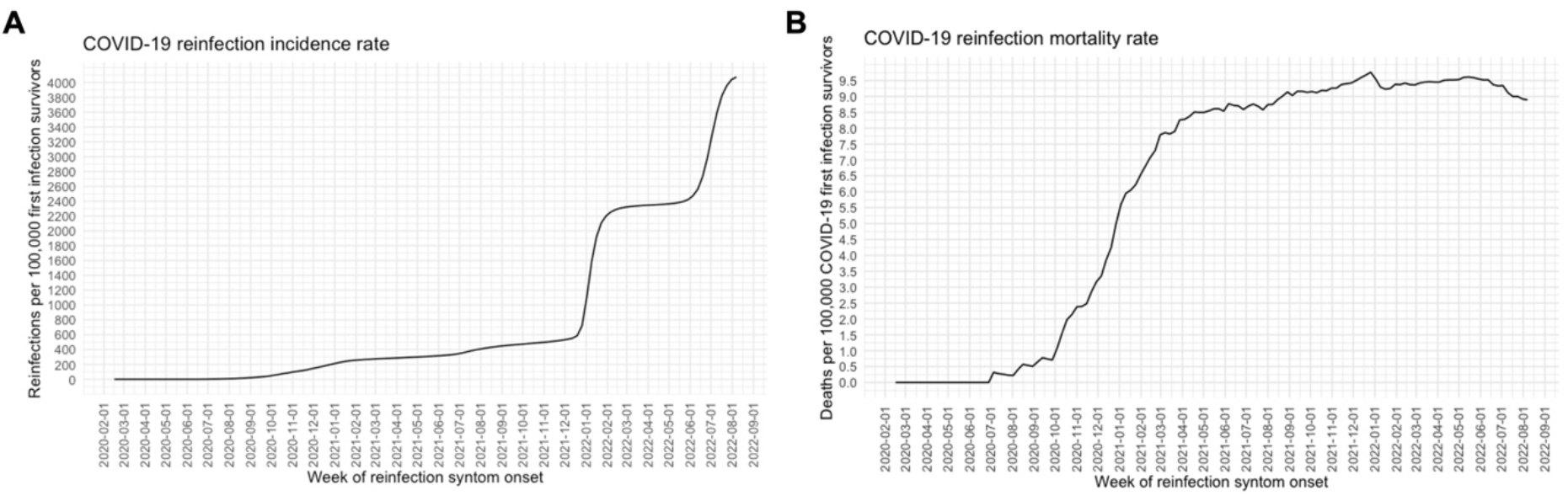
Incidence rates per 100,000 inhabitants of SARS-CoV-2 cases associated with reinfections (A) and reinfection-associated deaths (B) in survivors of first SARS-CoV-2 infections in Mexico from March 2020 until August 13th, 2022 (n=231,202).

### Risk of SARS-CoV-2 reinfection with hybrid immunity

We paired 158,609 cases of confirmed reinfection with 158,609 controls with primary infection and a second negative SARS-CoV-2 test ≥90 days after primary infection (**Supplementary Material**). Decreased risk of reinfection was associated with predominance of most SARS-CoV-2 variants compared to the ancestral strain, with higher protection observed for predominance of Gamma, Omicron BA.1 and BA.2 subvariants (**Figure 3**). Increased risk of reinfection was associated with predominance of the Omicron BA.4 and BA.5 subvariants, compared to periods of BA.1 subvariant predominance. Hospitalization during the first SARS-CoV-2 infection was associated with lower risk of reinfection, whilst having ≥6 months since the last immunity-generating event was associated with higher risk. The order of immunity-generating events did not largely impact the risk of reinfection compared to unvaccinated individuals with primary infection; however, the lowest risk was observed in fully boosted individuals prior to primary infection. A paradoxical increase in the risk of reinfection was observed in subjects fully vaccinated after primary infection; nevertheless, a higher risk in this category was observed for subjects with ≥6 months since last immunity-generating event. When stratifying models according to variant predominance at the time of reinfection, no significant changes were observed for these associations (**Supplementary Material**).

**Figure 3.**
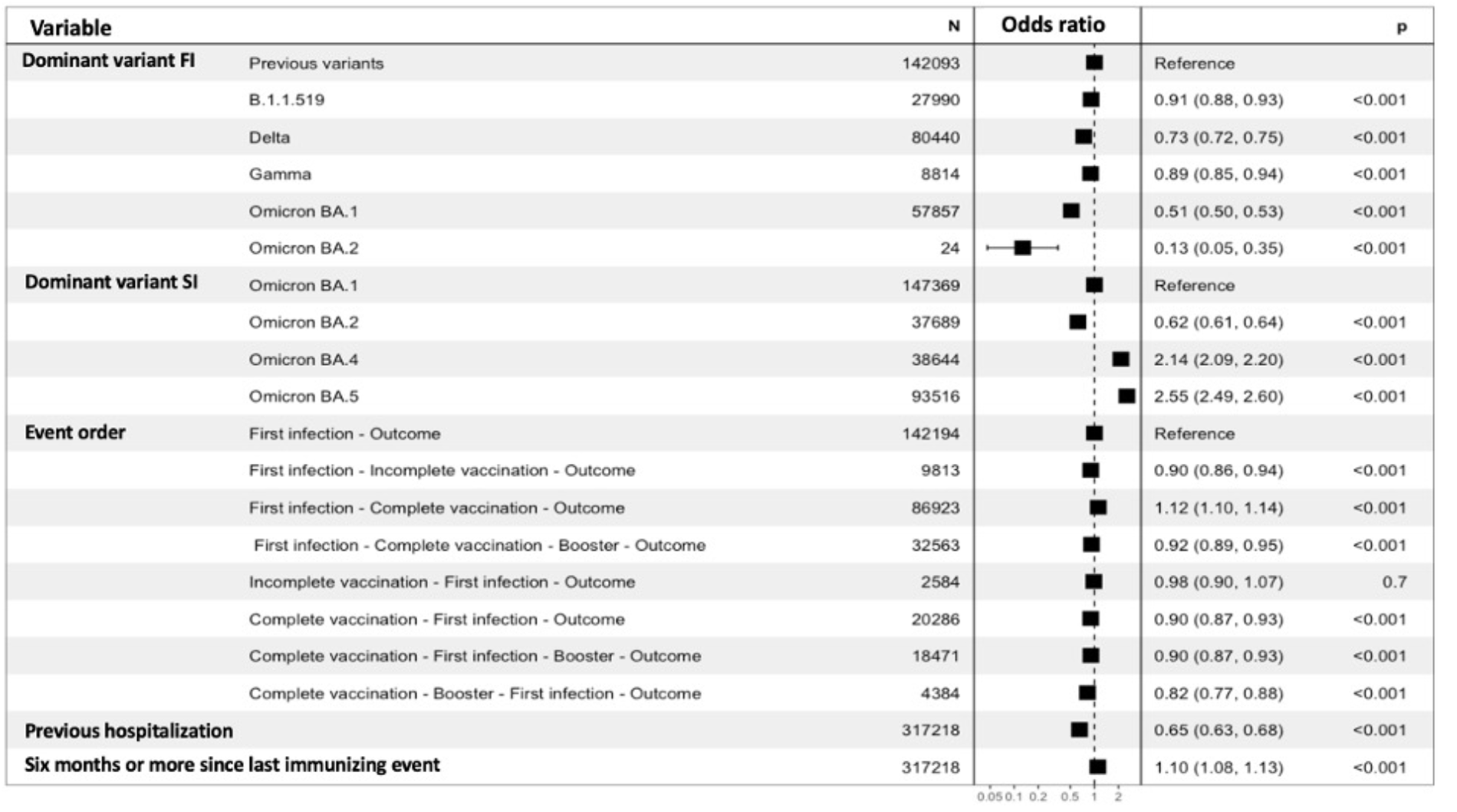
Conditional logistic regression model for risk of reinfection in subjects with confirmed reinfection (n=158,609) paired with subjects with first confirmed SARS-CoV-2 reinfection and a second negative test ≥90 days after the first infection (n= 158,609). Pairing was performed using propensity score matching for age and sex. **Abbreviations**: FI, first infection. SI, second infection.

### Risk of severe COVID-19 associated with reinfections

We explored risk factors in subjects with confirmed reinfection and severe COVID-19 (n=2,078) paired 1:4 with subjects with reinfection without severe COVID-19 (n=8,312) using propensity score matching for age and sex. Regarding primary infection, a decreased risk for severe COVID-19 was observed during periods of Gamma predominance compared to predominance of the ancestral strain, whilst an increased risk was associated with primary infection during periods of Omicron BA.1 subvariant predominance. A progressively decreased risk of severe COVID-19 was observed during periods of Omicron BA.2, BA.4 and BA.5 predominance, compared to reinfection during predominance of the BA.1 subvariant (**Figure 4**). Compared to unvaccinated individuals, all combination of immunity-generating events which included at least one vaccine dose were associated with >90% reduction in the risk of severe COVID-19, except for incomplete vaccination prior to primary infection, which yielded only a ∼87% reduction in risk. Having been hospitalized and survived during the primary SARS-CoV-2 infection was associated with higher risk of severe COVID-19 during reinfection, as well as having ≥6 months or more since the last immunity-generating event and having comorbid diabetes. Additional adjustment for age and sex to control for residual confounding did not modify these associations.

**Figure 4.**
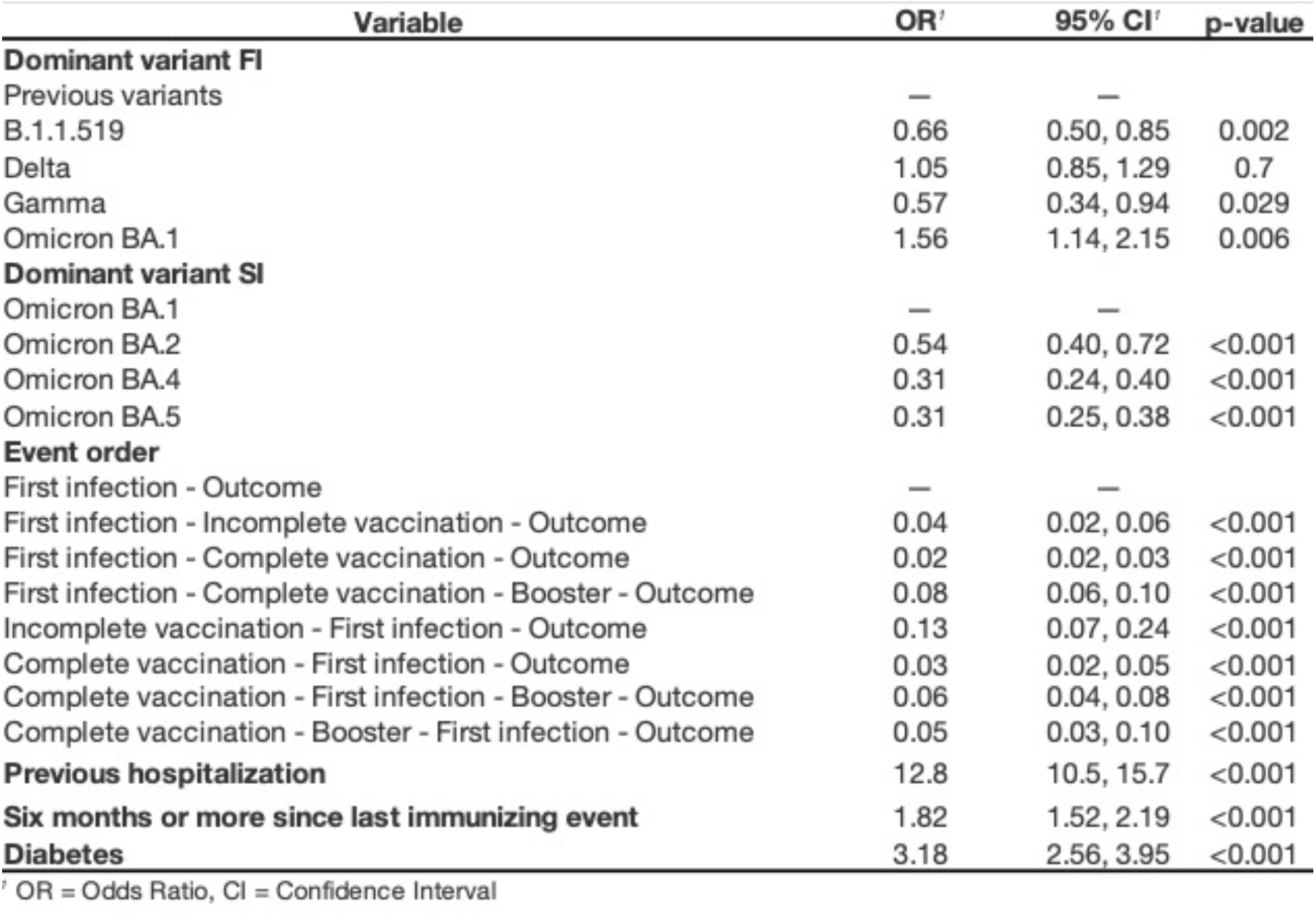
Conditional logistic regression model for risk of severe COVID-19, defined as an event of hospitalization, requirement for ICU admission or intubation and/or death in subjects with confirmed SARS-CoV-2 reinfection and severe COVID-19 (n=2,078) compared to mild SARS-CoV-2 reinfections (n=8,312), paired using propensity score matching for age and sex. **Abbreviations**: FI, first infection. SI, second infection.

### Homologous vs. heterologous boosters and risk of reinfection and severe outcomes

The most frequently reported booster shot in subjects evaluated for reinfection was heterologous vaccination of BNT162b2 boosted with ChAdOx1 (40.6%), followed by homologous vaccination with ChAdOx1 (23.5%), homologous vaccination with BNT162b2 (5.3%) and heterologous vaccination of Ad5-nCoV boosted with mRNA-12732 (4.8%, **Supplementary Material**). We observed 102,634 reinfections in fully vaccinated or boosted individuals with primary infection (n=183,986, 55.8%), amongst which 67,880 occurred in fully vaccinated individuals without boosting (n=121,727, 55.8%) and 34,754 occurred in individuals with boosters (n=62,259, 55.8%). Risk factors for SARS-CoV-2 reinfection included older age and female sex, whilst a higher risk was observed for reinfections which occurred in periods of Omicron BA.4 and BA.5 predominance (**Figure 5A**). Compared to fully vaccinated individuals without boosting, heterologous boosters were associated with ∼11% decreased risk of reinfection, with no significant difference for homologous boosters. Cases with ≥6 months since the last immunity-generating event had higher risk of reinfection. Increased risk of reinfection-associated severe COVID-19 was observed for older age whilst infection during predominance of Omicron BA.4 and BA.5 subvariants was associated with decreased risk (**Figure 5B**). Subjects with heterologous boosters and ≥6 months from the last immunity-generating event had ∼54% lower risk of severe COVID-19 compared to completely vaccinated individuals with ≥6 months from exposure; notably, no differences were observed for homologous boosters compared to subjects without boosting.

**Figure 5.**
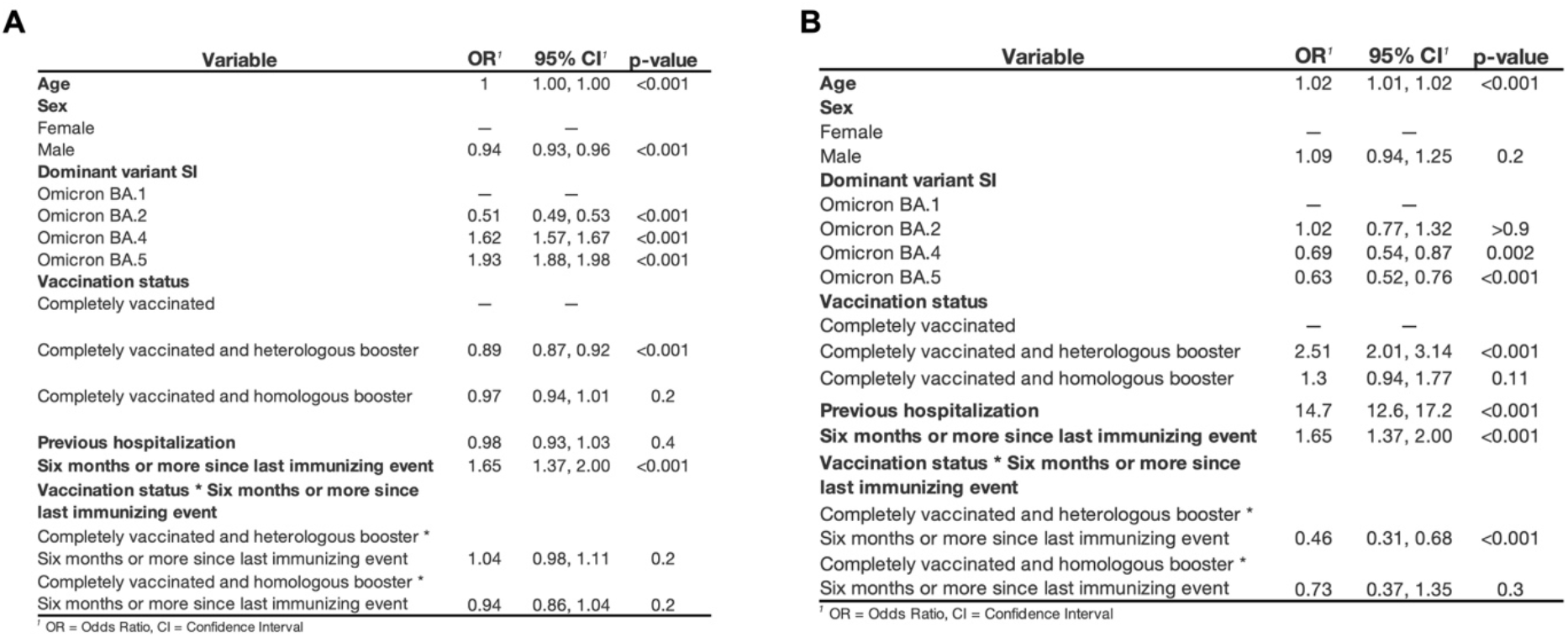
Conditional logistic regression model for risk of reinfection (A) and severe COVID-19 (B) in subjects with complete vaccination protocol (n=121,727) with or without an additional booster shot (n=62,259), to evaluate the impact of heterologous or homologous boosting in subjects with first confirmed SARS-CoV-2 infection. <u>**Abbreviations**</u>: SI, second infection.

## DISCUSSION

Here, we conducted one of the largest evaluations of the risk of reinfection and severe COVID-19 by analyzing 231,202 individuals with primary SARS-CoV-2 infection in Mexico, with 89.3% reinfections occurring during periods of Omicron predominance. In this setting, we were able to evaluate the role of hybrid immunity, severity of primary infection and influence of time-waning immunity on the risk of reinfection and reinfection-related severe COVID-19 risk. As previously reported, most reinfections were associated with Omicron and its subvariants (39,40), with rapid acceleration of reinfection rates during BA.1 and BA.5 subvariant predominance, and an intermediate plateau attributed to a stable primary infection-reinfection ratio. Reinfection-associated death rates were not modified by predominance of Omicron subvariants. Vaccination protected against reinfection, without significant influence from the order of immunity-generating events; nevertheless, the highest degree of protection was observed for fully vaccinated individuals boosted prior to primary infection, suggesting that in fully vaccinated naïve individuals, primary infection acts as an immune booster (41). For severe COVID-19, protection was conferred by primary infection with the dominant B.1.1.519 (42) or the Gamma variant, and was higher with Omicron BA.1 compared to subsequent subvariants. Hospitalization during primary infection >6 months since the last immunity-generating event and comorbid diabetes conferred a higher risk of severe COVID-19 reinfection. Finally, we observed a benefit of heterologous boosters over fully vaccinated individuals for protection against reinfection or severe COVID-19, confirming previous reports on the benefits of heterologous vaccination against COVID-19 (43,44).

Considering that approximately ∼50% of Mexican adults have antibodies against the Nucleocapsid protein of SARS-CoV-2 indicating exposure and likely a previous infection(45), and given the widespread circulation of Omicron, seroprevalence most likely increased along with the proportion of population at risk of reinfection (46). This was demonstrated by the increasing trends in the proportion of reinfections found in this study, representing as much as 12% of the weekly total COVID-19 cases (**Supplementary Material**). However, this proportion is likely higher due to limitations of case definitions and reporting. This proportion is expected to continue growing following the appearance of new variants and time-dependent waning of immune protection. The contribution of COVID-19 vaccines to hybrid immunity wanes within a few months of vaccination, particularly in the context of each new variant of concern (47); however, effectiveness is still maintained for Omicron and its subvariants, particularly against severe COVID-19. Therefore, vaccines are observed to still meet the strategic objectives highlighted in the WHO’s SAGE Roadmap for prioritizing uses of COVID-19 vaccines by preventing severe reinfections (48).

Amongst the strengths of our study, we highlight that it represents one of the largest reports of SARS-CoV-2 reinfections and risk factors in a country with high SARS-CoV-2 seroprevalence and large vaccination coverage. Furthermore, the use of a national epidemiological surveillance dataset of all suspected SARS-CoV-2 infections allows for adequate assessment of reinfections and its associated outcomes as well as the order of immunity-generating events to address the heterogeneity and complexity of hybrid immunity. Finally, given the large diversity of implemented vaccines in Mexico, we were also able to provide real-world evidence on the effectiveness of heterologous and homologous vaccine boosters in subjects with prior SARS-CoV-2 infection, which have been scarcely reported. Amongst the limitations to be acknowledged are the definition of reinfection, with recent evidence demonstrating that reinfections can occur over a shorter period, and may incorrectly exclude some reinfections in our study (49). Furthermore, when analyzing risk associated with SARS-CoV-2 variants, we assumed that infections were caused by predominant variants during each period; nevertheless, this type of inference has been used as an approach to variant analysis when genomic data is not available (10,38,42). This approach limits our capacity to differentiate the effect of high community transmission pressure on reinfections from the ability of variants to evade hybrid immunity. However, higher transmissibility of new SARS-CoV-2 variants is intimately related to their virulence and should therefore be considered a consequence of the later. Finally, we did not perform individual analyses for each vaccine and booster combination, provided that not all combinations had large enough number of outcomes to allow for adequate comparisons. The role of individual vaccination and boosters and their interaction with predominant circulating variants remains as an area of opportunity for future studies. The frequency in which booster shots will need to be applied in the future is still uncertain.

Given that most reinfections occurred during periods of Omicron predominance and that immunity boosting by infection with Omicron seems to be low, further evaluations are required to explore the role of second boosters, subsequent infections, and the order of these immunity-generating events in modifying the risk of reinfection and reinfection-associated severe COVID-19 as the pandemic further progresses and new SARS-CoV-2 variants continue to emerge. However, as of now, vaccination continues to be our strongest defense against COVID-19 and should therefore be prioritized in those unvaccinated or those who have not received their first booster shot to prevent sever outcomes. Supported by the evidence of no difference in severity between infections and reinfections, the high prevalence of cardio-metabolic diseases in Mexico and the association found in our study between both diabetes and hospitalization during first infection with severe reinfections, further efforts should be made on increasing primary vaccination and booster rates to reduce the risk of SARS-CoV-2 reinfection, severe-COVID-19, and risk of post-acute COVID-19 syndrome associated with reinfection, particularly for variants with increased transmission and associated immune evasion such as Omicron (50). As reinfections become more frequent and time between them keeps on shortening (51), surveillance systems may benefit from the study of SARS-CoV-2 reinfections.

## Data Availability

DATA AVAILABILITY: All codes are available to assess statistical analysis at https://github.com/oyaxbell/reinfections_covid/. Data is available upon request to the General Directorate of Epidemiology of the Mexican Ministry of Health.

## ETHICS APPROVAL

This project was registered and approved by the Ethics and Research Committee at Instituto Nacional de Geriatría, project number DI-PI-005/2021.

## DATA AVAILABILITY

All codes are available to assess statistical analysis at https://github.com/oyaxbell/reinfections_covid/. Data is available upon request to the General Directorate of Epidemiology of the Mexican Ministry of Health.

## AUTHOR CONTRIBUTIONS

Research idea and study design: JAMG, OYBC; data acquisition: JAMG, CAZJ, RIGV, GGR, HLG; data analysis/interpretation: JAMG, OYBC, NEAV, CAFM, DRG, AVV; statistical analysis: JAMG, OYBC; manuscript drafting: JAMG, OYBC, NEAV, CAFM, DRG, AVV, SIVF; supervision or mentorship: OYBC. Each author contributed important intellectual content during manuscript drafting or revision and accepted accountability for the overall work by ensuring that questions pertaining to the accuracy or integrity of any portion of the work are appropriately investigated and resolved.

## ACKNOWLEDGMENTS

NEAV, AVV and CAFM are enrolled at the PECEM program of the Faculty of Medicine at UNAM. NEAV is supported by CONACyT. The authors would like to acknowledge the invaluable work of all of Mexico’s healthcare community in managing the COVID-19 pandemic. Their participation in the COVID-19 surveillance program along with open data sharing has made this work a reality, we are thankful for your effort.

