## Supplementary Material for "Protection of hybrid immunity against SARS-CoV-2 reinfection and severe COVID-19 during periods of Omicron variant predominance in Mexico"

**SUPPLEMENTARY TABLES**

**TABLES**

**Supplementary Table 1.** Socio-demographic and clinical characteristics of SARS-CoV-2 reinfections in Mexico from March 2020 until August 13^th^, 2022

| **Category** | **Variable** | **N** | **(%)** |
| --- | --- | --- | --- |
|  | SARS-CoV-2 Reinfections | 231,202 | 100.0 |
| **Sex** |  |  |  |
|  | Male | 92,210 | 39.88 |
|  | Female | 138,992 | 60.12 |
| **Age group** |  |  |  |
|  | 0-10 | 1,690 | 0.73 |
|  | 11-20 | 6,850 | 2.96 |
|  | 21-30 | 56,463 | 24.42 |
|  | 31-40 | 69,952 | 30.26 |
|  | 41-50 | 56,184 | 24.30 |
|  | 51-60 | 29,546 | 12.78 |
|  | 61-70 | 7,961 | 3.44 |
|  | 71-80 | 2,057 | 0.89 |
|  | 81-90 | 431 | 0.19 |
|  | 91 and more | 68 | 0.03 |
| **Vaccination** |  |  |  |
| *Immunity-generating event order* | |  |  |
|  | First infection - Reinfection | 109,936 | 47.55 |
|  | First infection - Complete vaccination - Reinfection | 65,145 | 28.18 |
|  | First infection - Complete vaccination - Booster - Reinfection | 22,492 | 9.73 |
|  | Complete vaccination - First infection - Reinfection | 11,989 | 5.19 |
|  | Complete vaccination - First infection - Booster - Reinfection | 10,585 | 4.58 |
|  | First infection - Incomplete vaccination - Reinfection | 7,192 | 3.11 |
|  | Complete vaccination - Booster - First infection - Reinfection | 2,300 | 0.99 |
|  | Incomplete vaccination - First infection - Reinfection | 1,563 | 0.68 |
| *Vaccine platform for full vaccine protocol (n=117,941)* | |  |  |
|  | mRNA | 49,042 | 41.58 |
|  | Inactivated pathogen vaccine | 6,906 | 5.86 |
|  | Protein subunit vaccine | 55 | 0.05 |
|  | Replication defective viral vector | 61,938 | 52.52 |
| Booster platform (n= 34,784) | |  |  |
|  | mRNA | 4,353 | 12.51 |
|  | Inactivated pathogen vaccine | 239 | 0.69 |
|  | Protein subunit vaccine | 19 | 0.05 |
|  | Replication defective viral vector | 30,173 | 86.74 |
| Booster schedule (n= 34,784) | |  |  |
|  | Heterologous | 22,595 | 64.96 |
|  | Homologous | 12,023 | 34.56 |
| **Comorbidities** |  |  |  |
|  | Diabetes | 13,803 | 5.97 |
|  | Obesity | 19,276 | 8.34 |
| **SARS-CoV-2 dominant variant circulating at the time of infection** | | |  |
| First infection |  |  |  |
|  | Ancestral variants | 115,731 | 50.06 |
|  | B.1.1.519 | 21,603 | 9.34 |
|  | Delta | 53,419 | 23.10 |
|  | Gamma | 6,528 | 2.82 |
|  | Omicron BA.1 | 33,911 | 14.67 |
|  | Omicron BA.2 | 10 | 0.00 |
| Reinfection |  |  |  |
|  | Ancestral variants | 3,394 | 1.47 |
|  | B.1.1.519 | 1,986 | 0.86 |
|  | Delta | 17,435 | 7.54 |
|  | Gamma | 764 | 0.33 |
|  | Omicron BA.1 | 84,949 | 36.74 |
|  | Omicron BA.2 | 14,789 | 6.40 |
|  | Omicron BA.4 | 30,049 | 13.00 |
|  | Omicron BA.5 | 77,836 | 33.67 |
| **Time to reinfection (Median,Q1,Q3)** 362 days (196,531) | | | |
| **Severe outcomes** | |  |  |
|  | Hospitalization | 3,261 | 1.41 |
|  | Intubation | 122 | 0.05 |
|  | ICU admission | 130 | 0.06 |
|  | Death | 515 | 0.22 |

**Supplementary Table 2.** Demographic and clinical characteristics of cases with confirmed SARS-CoV-2 reinfection with subjects with a second negative SARS-CoV-2 test during this period paired for age and sex who acquired reinfection during periods of Omicron variant predominance in Mexico.

|  | **1:1 ratio** | **Cases (n = 158,609)** | | | | **Controls (n = 158,609)** | | | |  | **P** |
| --- | --- | --- | --- | --- | --- | --- | --- | --- | --- | --- | --- |
|  | **Variable** | **Categories (%)** | | | | **Categories (%)** | | | |  |  |
| **Sex** |  |  |  |  |  |  |  |  |  |  |  |
|  | Male | 43,281 | ( | 27.29 | ) | 65,651 | ( | 41.39 | ) | < | 0.000 |
|  | Female | 115,328 | ( | 72.71 | ) | 92,958 | ( | 58.61 | ) |  |  |
| **Age group** |  |  |  |  |  |  |  |  |  |  |  |
|  | Median | 38 |  |  |  | 37 |  |  |  | < | 0.000 |
|  | Q1 | 32 |  |  |  | 28 |  |  |  |  |  |
|  | Q3 | 46 |  |  |  | 48 |  |  |  |  |  |
| **Vaccination** |  |  |  |  |  |  |  |  |  |  |  |
| Event order |  |  |  |  |  |  |  |  |  |  |  |
|  | First infection - Outcome | 71,679 | ( | 45.19 | ) | 70,515 | ( | 44.46 | ) | < | 0.000 |
|  | First infection - Complete vaccination - Outcome | 44,750 | ( | 28.21 | ) | 42,173 | ( | 26.59 | ) |  |  |
|  | First infection - Complete vaccination - Booster - Outcome | 17,604 | ( | 11.10 | ) | 14,959 | ( | 9.43 | ) |  |  |
|  | Complete vaccination - First infection - Outcome | 8,749 | ( | 5.52 | ) | 11,537 | ( | 7.27 | ) |  |  |
|  | Complete vaccination - First infection - Booster - Outcome | 8,361 | ( | 5.27 | ) | 10,110 | ( | 6.37 | ) |  |  |
|  | First infection - Incomplete vaccination - Outcome | 4,477 | ( | 2.82 | ) | 5,336 | ( | 3.36 | ) |  |  |
|  | Complete vaccination - Booster - First infection - Outcome | 1,811 | ( | 1.14 | ) | 2,573 | ( | 1.62 | ) |  |  |
|  | Incomplete vaccination - First infection - Outcome | 1,178 | ( | 0.74 | ) | 1,406 | ( | 0.89 | ) |  |  |
| Vaccine platform | | **n = 86,930** | |  |  | **n = 88,094** | |  |  |  |  |
|  | mRNA | 35,529 | ( | 40.87 | ) | 33,843 | ( | 38.42 | ) | < | 0.000 |
|  | Inactivated pathogen vaccine | 4,621 | ( | 5.32 | ) | 4,698 | ( | 5.33 | ) |  |  |
|  | Protein subunit vaccine | 41 | ( | 0.05 | ) | 40 | ( | 0.05 | ) |  |  |
|  | Replication defective viral vector | 46,739 | ( | 53.77 | ) | 49,513 | ( | 56.20 | ) |  |  |
| Booster platform | | **n = 27,611** | |  |  | **n = 27,505** | |  |  |  |  |
|  | mRNA | 3,554 | ( | 12.87 | ) | 3,994 | ( | 14.52 | ) |  | 0.0000001 |
|  | Inactivated pathogen vaccine | 169 | ( | 0.61 | ) | 204 | ( | 0.74 | ) |  |  |
|  | Protein subunit vaccine | 17 | ( | 0.06 | ) | 16 | ( | 0.06 | ) |  |  |
|  | Replication defective viral vector | 23,871 | ( | 86.45 | ) | 23,291 | ( | 84.68 | ) |  |  |
| Booster scheme | |  |  |  |  |  |  |  |  |  |  |
|  | Heterologous | 18,148 | ( | 65.73 | ) | 18,514 | ( | 67.31 | ) |  | 0.0000001 |
|  | Homologous | 9,463 | ( | 34.27 | ) | 8,991 | ( | 32.69 | ) |  |  |
| **Comorbidities** | |  |  |  |  |  |  |  |  |  |  |
|  | Diabetes | 8,197 | ( | 5.17 | ) | 10,585 | ( | 6.67 | ) | < | 0.001 |
|  | Obesity | 13,801 | ( | 8.70 | ) | 11,021 | ( | 6.95 | ) | < | 0.001 |
| **SARS-CoV-2 dominant variant circulating at the time of infection** | | |  |  |  |  |  |  |  |  |  |
| First infection | |  |  |  |  |  |  |  |  |  |  |
|  | Ancestral strains | 76,351 | ( | 48.14 | ) | 65,742 | ( | 41.45 | ) | < | 0.001 |
|  | B.1.1.519 | 14,397 | ( | 9.08 | ) | 13,593 | ( | 8.57 | ) |  |  |
|  | Delta | 37,894 | ( | 23.89 | ) | 42,546 | ( | 26.82 | ) |  |  |
|  | Gamma | 4,491 | ( | 2.83 | ) | 4,323 | ( | 2.73 | ) |  |  |
|  | Omicron BA.1 | 25,471 | ( | 16.06 | ) | 32,386 | ( | 20.42 | ) |  |  |
|  | Omicron BA.2 | 5 | ( | 0.00 | ) | 19 | ( | 0.01 | ) |  |  |
| Reinfection |  |  |  |  |  |  |  |  |  |  |  |
|  | Omicron BA.1 | 67,406 | ( | 42.50 | ) | 79,963 | ( | 50.42 | ) | < | 0.001 |
|  | Omicron BA.2 | 10,819 | ( | 6.82 | ) | 26,870 | ( | 16.94 | ) |  |  |
|  | Omicron BA.4 | 22,446 | ( | 14.15 | ) | 16,198 | ( | 10.21 | ) |  |  |
|  | Omicron BA.5 | 57,938 | ( | 36.53 | ) | 35,578 | ( | 22.43 | ) |  |  |
| **Previous outcomes** | |  |  |  |  |  |  |  |  |  |  |
|  | Hospitalized during first infection | 4,727 | ( | 2.98 | ) | 6,509 | ( | 4.10 | ) | < | 0.001 |
| **Time since last immunizing event** | |  |  |  |  |  |  |  |  |  |  |
|  | 6 months or more | 91,277 | ( | 57.55 | ) | 76,279 | ( | 48.09 | ) | < | 0.001 |
|  | < 6 months | 67,332 | ( | 42.45 | ) | 82,330 | ( | 51.91 | ) |  |  |

**Supplementary Table 3.** Demographic and clinical characteristics of cases with confirmed SARS-CoV-2 reinfection and severe COVID-19 with cases with mild SARS-CoV-2 reinfection during this period paired for age and sex in a 1:4 ratio.

|  | **1:4 ratio** | **Cases (n = 2,078)** | | | | **Controls (n = 8,312)** | | | | **P** | |
| --- | --- | --- | --- | --- | --- | --- | --- | --- | --- | --- | --- |
|  | **Variable** | **Categories (%)** | | | | **Categories (%)** | | | |  | |
|  |  |  | ## | ##### |  |  |  | ##### |  |  |  |
| **Sex** |  |  |  |  |  |  |  |  |  |  |  |
|  | Male | 937 | ( | 45.09 | ) | 3,745 | ( | 45.06 | ) |  | 0.97 |
|  | Female | 1,141 | ( | 54.91 | ) | 4,567 | ( | 54.94 | ) |  |  |
| **Age** |  |  |  |  |  |  |  |  |  |  |  |
|  | Median | 43 |  |  |  | 43 |  |  |  |  | 0.5952 |
|  | Q1 | 31 |  |  |  | 31 |  |  |  |  |  |
|  | Q3 | 58 |  |  |  | 57 |  |  |  |  |  |
| **Vaccination** |  |  |  |  |  |  |  |  |  |  |  |
| Event order |  |  |  |  |  |  |  |  |  |  |  |
|  | First infection - Outcome | 1,193 | ( | 57.41 | ) | 875 | ( | 10.53 | ) | < | 0.001 |
|  | First infection - Complete vaccination - Outcome | 395 | ( | 19.01 | ) | 3,729 | ( | 44.86 | ) |  |  |
|  | First infection - Complete vaccination - Booster - Outcome | 220 | ( | 10.59 | ) | 1,556 | ( | 18.72 | ) |  |  |
|  | Complete vaccination - First infection - Outcome | 92 | ( | 4.43 | ) | 875 | ( | 10.53 | ) |  |  |
|  | Complete vaccination - First infection - Booster - Outcome | 96 | ( | 4.62 | ) | 751 | ( | 9.04 | ) |  |  |
|  | First infection - Incomplete vaccination - Outcome | 41 | ( | 1.97 | ) | 285 | ( | 3.43 | ) |  |  |
|  | Complete vaccination - Booster - First infection - Outcome | 20 | ( | 0.96 | ) | 175 | ( | 2.11 | ) |  |  |
|  | Incomplete vaccination - First infection - Outcome | 21 | ( | 1.01 | ) | 66 | ( | 0.79 | ) |  |  |
| Vaccine platform |  | **n=885** | | |  | **n=7,437** | | |  |  |  |
|  | mRNA | 549 | ( | 62.03 | ) | 267 | ( | 3.59 | ) | < | 0.001 |
|  | Inactivated pathogen vaccine | 33 | ( | 3.73 | ) | 55 | ( | 0.74 | ) |  |  |
|  | Protein subunit vaccine | 1 | ( | 0.11 | ) | 0 | ( | 0.00 | ) |  |  |
|  | Replication defective viral vector | 302 | ( | 34.12 | ) | 7,115 | ( | 95.67 | ) |  |  |
| Booster platform |  | **n=334** | | |  | **n=2,465** | | |  |  |  |
|  | mRNA | 28 | ( | 8.38 | ) | 354 | ( | 14.36 | ) |  | 0.00038 |
|  | Inactivated pathogen vaccine | 7 | ( | 2.10 | ) | 14 | ( | 0.57 | ) |  |  |
|  | Protein subunit vaccine | 0 | ( | 0.00 | ) | 1 | ( | 0.04 | ) |  |  |
|  | Replication defective viral vector | 299 | ( | 89.52 | ) | 2,096 | ( | 85.03 | ) |  |  |
| Booster scheme |  |  |  |  |  |  |  |  |  |  |  |
|  | Heterologous | 259 | ( | 77.54 | ) | 1,650 | ( | 66.94 | ) |  | 0.00009 |
|  | Homologous | 75 | ( | 22.46 | ) | 815 | ( | 33.06 | ) |  |  |
| **Comorbidities** |  |  |  |  |  |  |  |  |  |  |  |
|  | Diabetes | 416 | ( | 20.02 | ) | 777 | ( | 9.35 | ) | < | 0.001 |
|  | Obesity | 194 | ( | 9.34 | ) | 821 | ( | 9.88 | ) |  | 0.457 |
| **SARS-CoV-2 dominant variant circulating at the time of infection** | | |  |  |  |  |  |  |  |  |  |
| First infection |  |  |  |  |  |  |  |  |  |  |  |
|  | Previously circulating variants | 1,075 | ( | 51.73 | ) | 3,768 | ( | 45.33 | ) |  | 0.001 |
|  | B.1.1.519 | 168 | ( | 8.08 | ) | 881 | ( | 10.60 | ) |  |  |
|  | Delta | 510 | ( | 24.54 | ) | 2,106 | ( | 25.34 | ) |  |  |
|  | Gamma | 37 | ( | 1.78 | ) | 237 | ( | 2.85 | ) |  |  |
|  | Omicron BA.1 | 288 | ( | 13.86 | ) | 1,320 | ( | 15.88 | ) |  |  |
| Reinfection |  |  |  |  |  |  |  |  |  |  |  |
|  | Omicron BA.1 | 1,075 | ( | 51.73 | ) | 3,242 | ( | 39.00 | ) | < | 0.000 |
|  | Omicron BA.2 | 167 | ( | 8.04 | ) | 659 | ( | 7.93 | ) |  |  |
|  | Omicron BA.4 | 220 | ( | 10.59 | ) | 1,306 | ( | 15.71 | ) |  |  |
|  | Omicron BA.5 | 616 | ( | 29.64 | ) | 3,105 | ( | 37.36 | ) |  |  |
| **Previous outcomes** | |  |  |  |  |  |  |  |  |  |  |
|  | Hospitalized during first infection | 751 | ( | 36.14 | ) | 403 | ( | 4.85 | ) | < | 0.000 |
| **Time since last immunizing event** | |  |  |  |  |  |  |  |  |  |  |
|  | 6 months or more | 1,312 | ( | 63.14 | ) | 3,330 | ( | 40.06 | ) | < | 0.000 |
|  | < 6 months | 766 | ( | 36.86 | ) | 4,982 | ( | 59.94 | ) |  |  |

**SUPPLEMENTARY FIGURES**

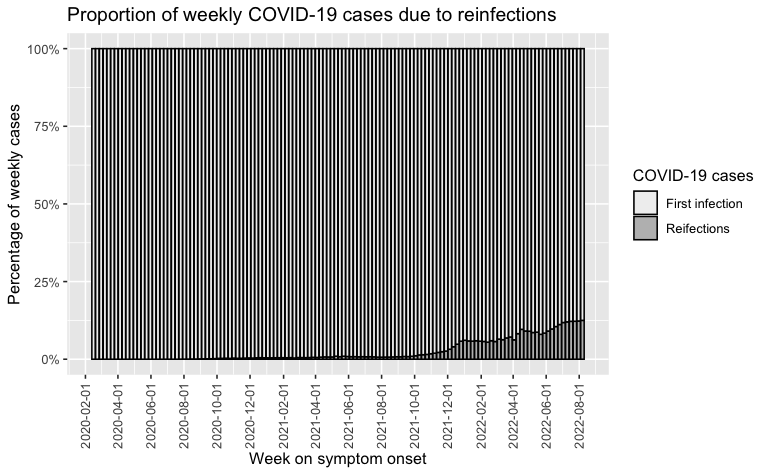

**Supplementary Figure 1.** Proportion of weekly cases attributable to SARS-CoV-2 reinfections from March 2020 until August 13^th^, 2022.

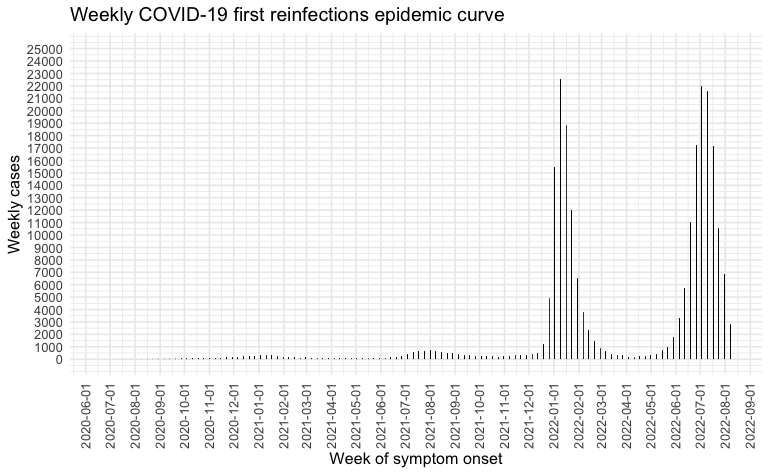

**Supplementary Figure 2.** Epidemic curve of weekly SARS-CoV-2 reinfections registered in Mexico during the study period according to week of symptom onset.

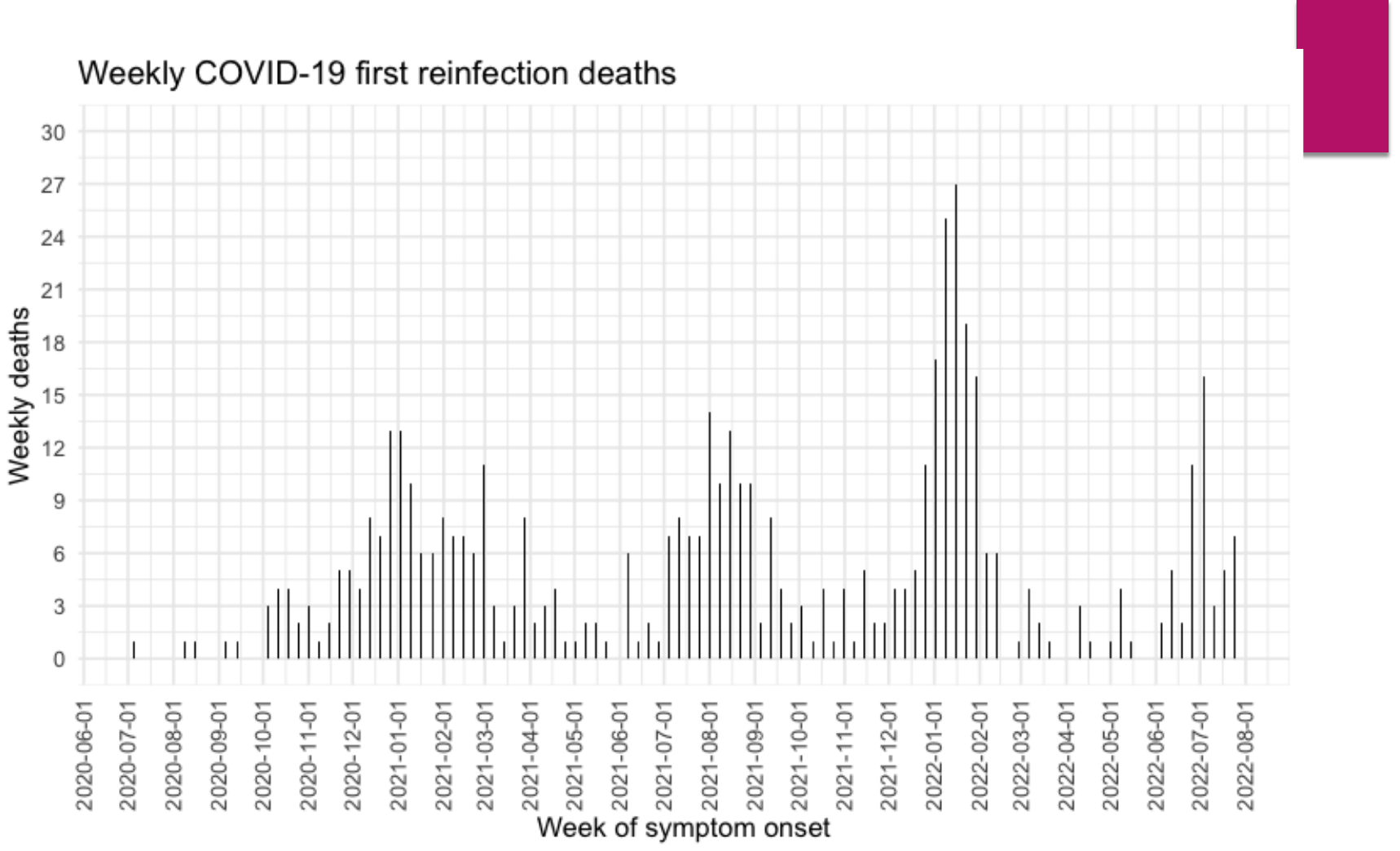

**Supplementary Figure 3.** Epidemic curve of weekly deaths associated with SARS-CoV-2 reinfections registered in Mexico during the study period according to week of symptom onset.

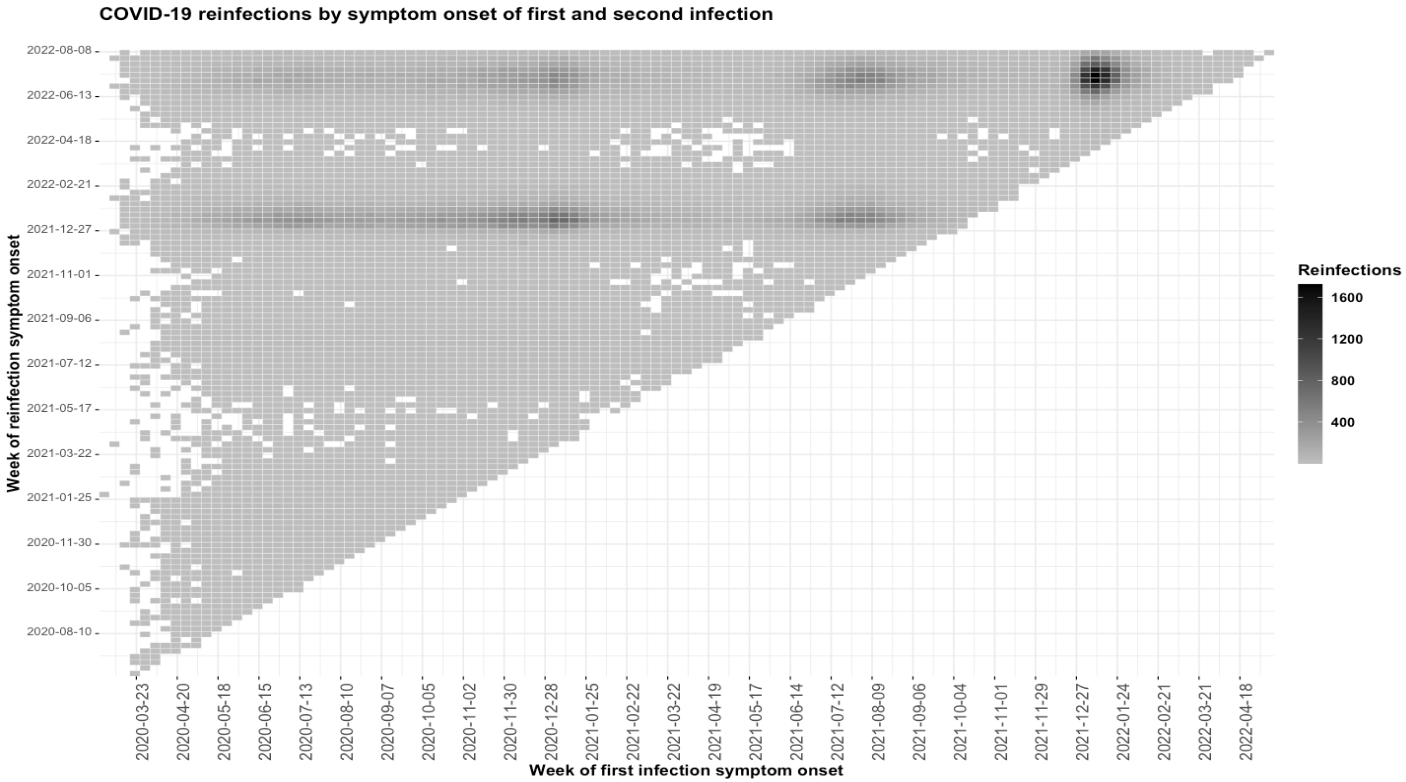

**Supplementary Figure 4.** Matrix of first SARS-CoV-2 infections and reinfections according to week of symptom onset in Mexico during the study period.

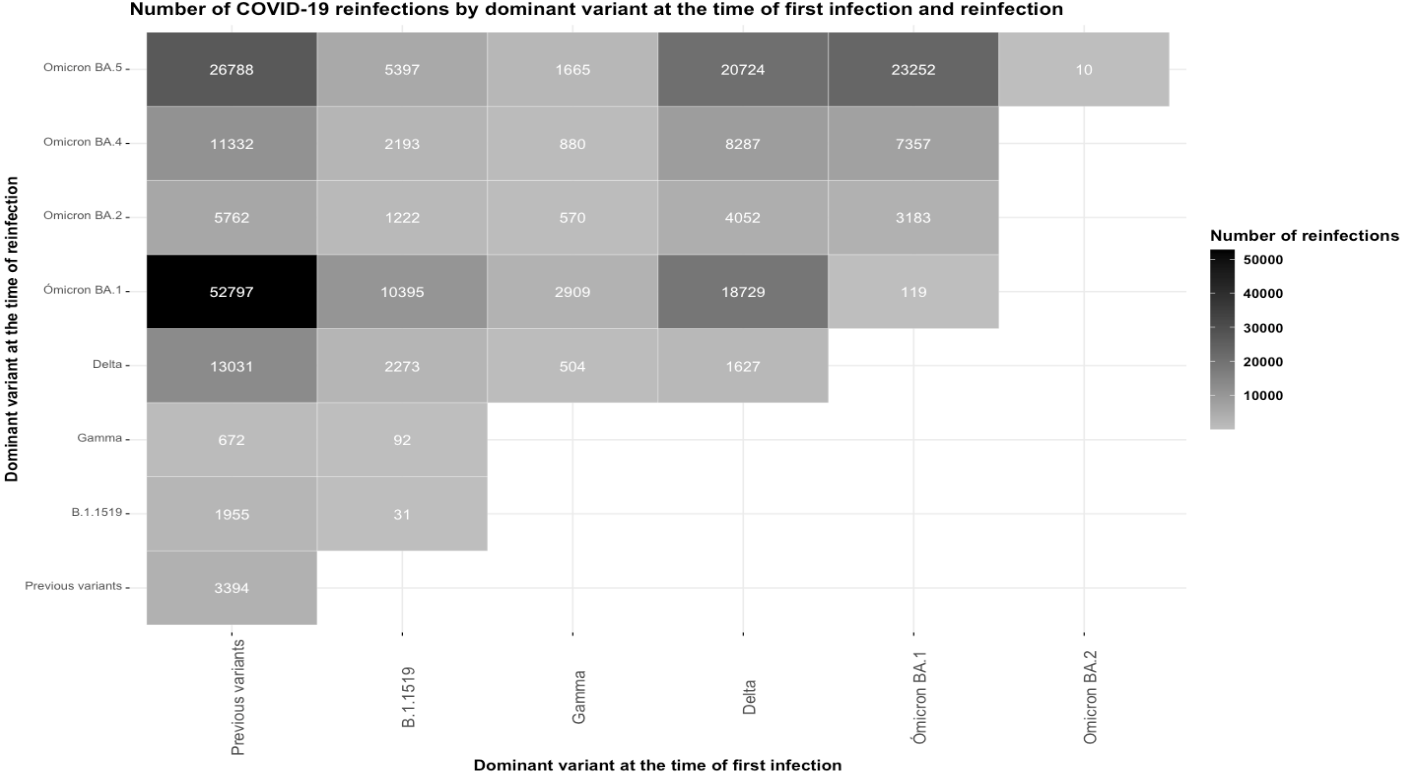

**Supplementary Figure 5.** Matrix of SARS-CoV-2 reinfections according to dominant variant at the time of first and second SARS-CoV-2 infection in Mexico during the study period.

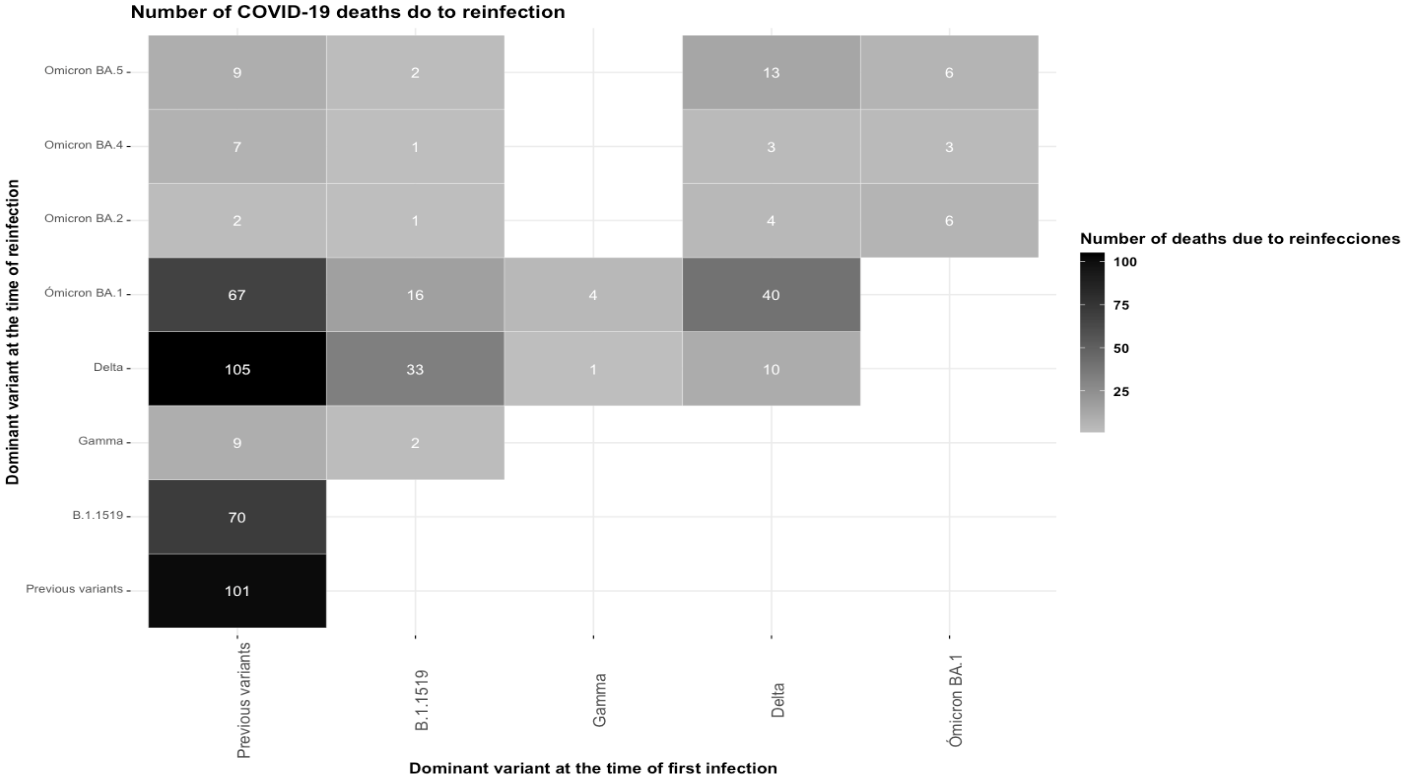

**Supplementary Figure 6.** Matrix of deaths associated to reinfections according to dominant variant at the time of first and second SARS-CoV-2 infection in Mexico during the study period.

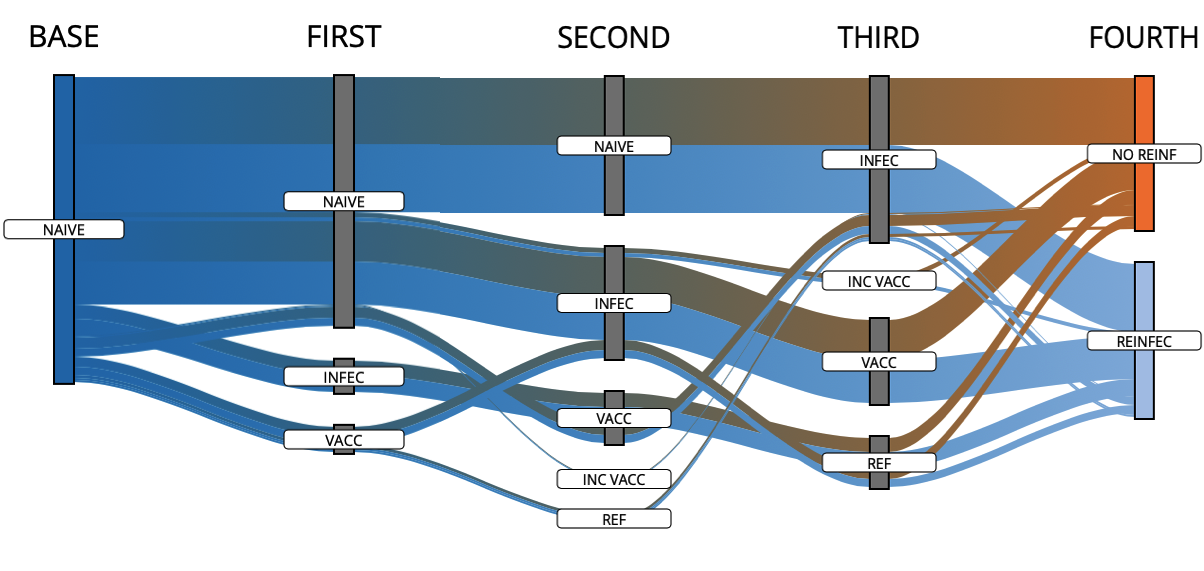

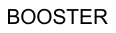

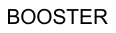

**Supplementary Figure 7.** Flow of participants according to the order of immunity-generating events in cases with primary SARS-CoV-2 infection and reinfection (n=158,609) compared to controls (n=158,609) with primary infection and a second negative RT-PCR or antigen test ≥90 days from the date of symptom onset or primary infection. **Abbreviations:** NAÏVE, individuals without primary SARS-CoV-2 infection; INFEC, Primary SARS-CoV-2 infection; VACC, fully vaccinated, INC VACC, incompletely vaccinated; BOOSTER, vaccine booster shot; NO REINF, No reinfection; REINFEC, SARS-CoV-2 reinfection

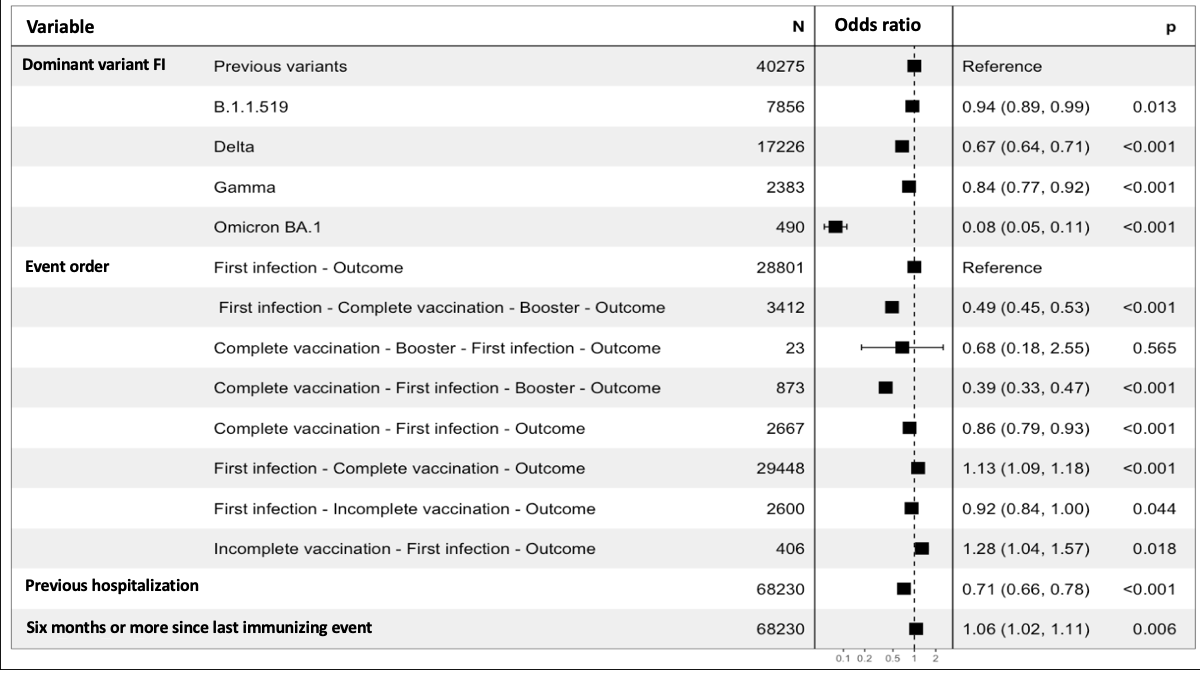

**Supplementary Figure 8.** Risk of reinfection during periods of predominance of the Omicron BA.1 subvariant in Mexico compared to subjects with a second negative SARS-CoV-2 test during this period paired for age and sex. **Abbreviations:** FI, first infection,

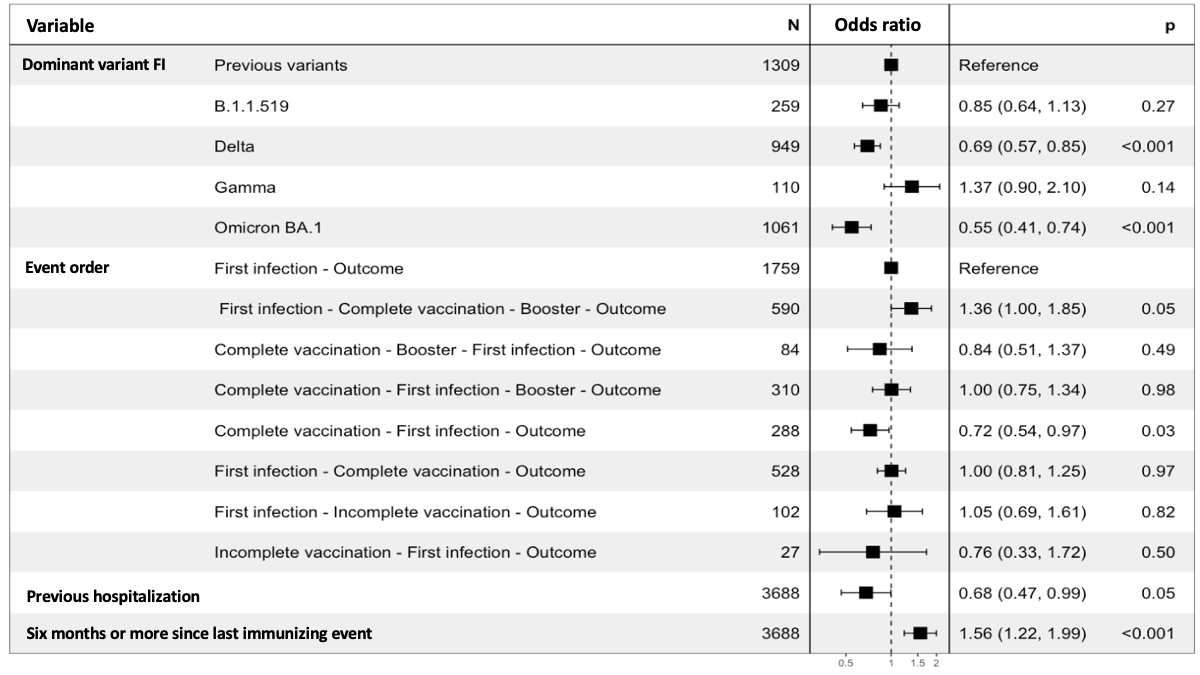

**Supplementary Figure 8.** Risk of reinfection during periods of predominance of the Omicron BA.2 subvariant in Mexico compared to subjects with a second negative SARS-CoV-2 test during this period paired for age and sex. **Abbreviations:** FI, first infection,

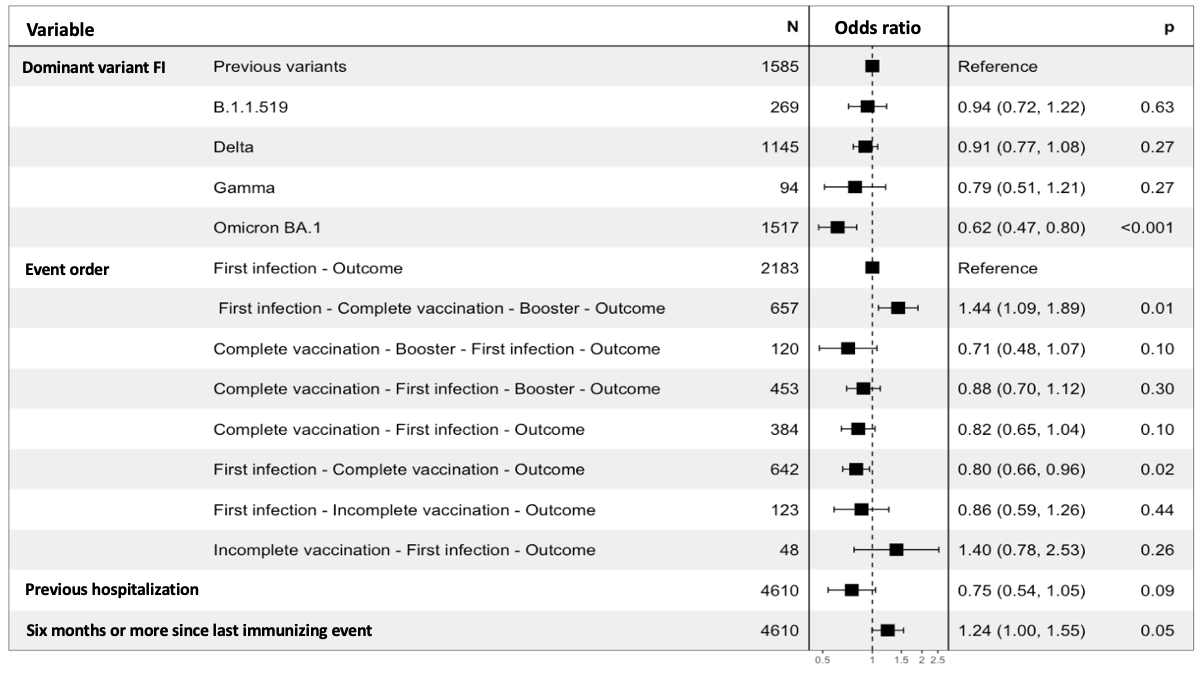

**Supplementary Figure 9.** Risk of reinfection during periods of predominance of the Omicron BA.4 subvariant in Mexico compared to subjects with a second negative SARS-CoV-2 test during this period paired for age and sex. **Abbreviations:** FI, first infection,

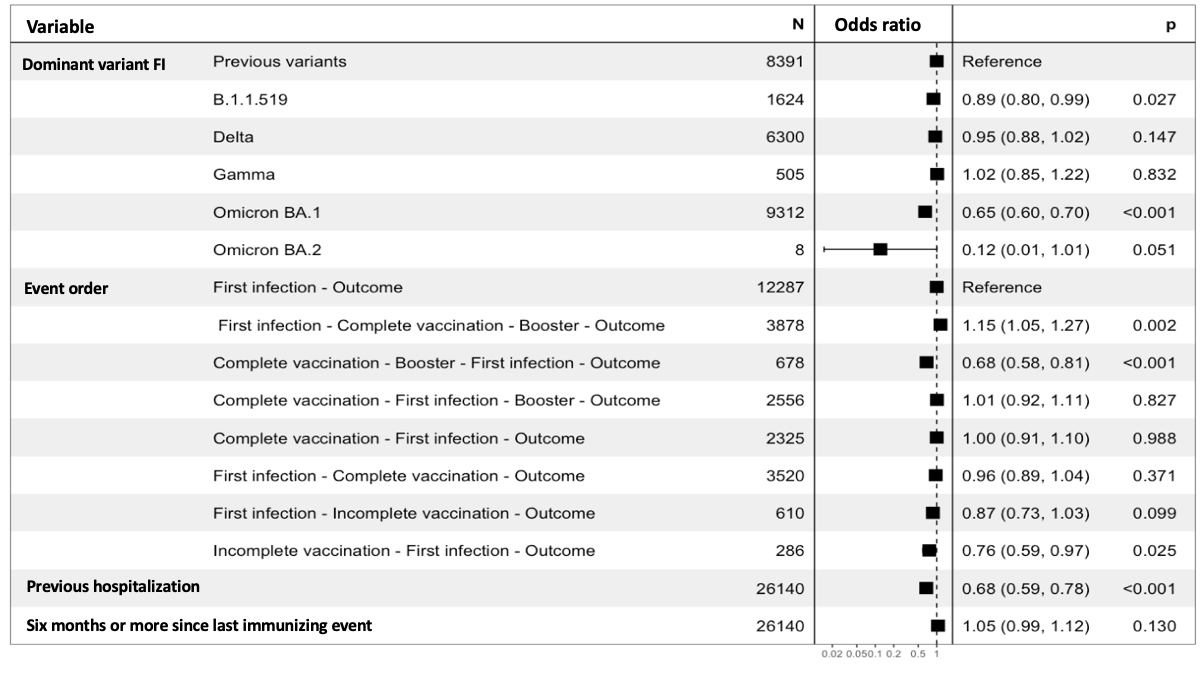

**Supplementary Figure 10.** Risk of reinfection during periods of predominance of the Omicron BA.5 subvariant in Mexico compared to subjects with a second negative SARS-CoV-2 test during this period paired for age and sex. **Abbreviations:** FI, first infection,

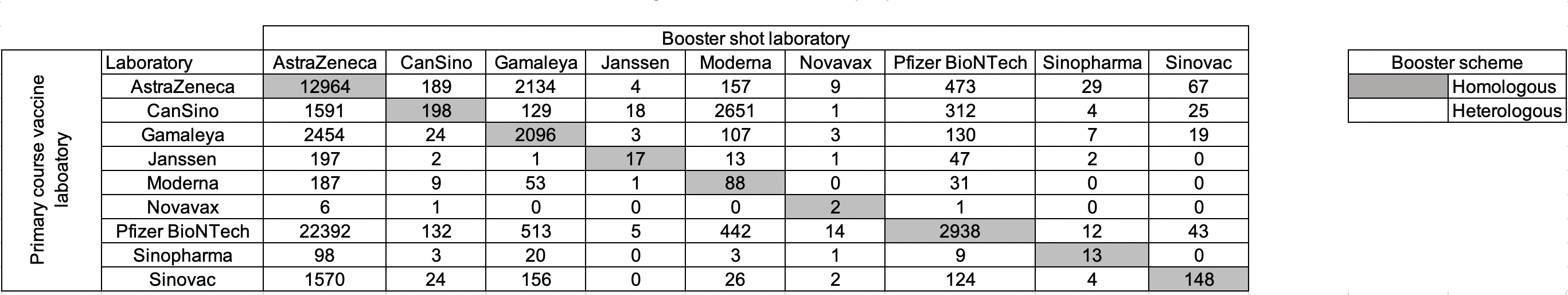

**Supplementary Figure 11.** Matrix of booster shot combinations in subjects with reinfections and paired subjects with a second negative SARS-CoV-2 test ≥90 days after the first infection in Mexico from March 3^rd^, 2020, until August 13^th^, 2022 further categorized by homologous compared to heterologous schemes.
